# Determination of Antibiotic Resistant Bacteria and Antibiotic Residues in Red Meat

**DOI:** 10.64898/2026.08.10.26360071

**Authors:** Nishe Saha, Sharmin afroz, Kallyanmoy Das, Md. Rimon Bhuiyan, Anna Purnna Ray, Md. Ahsan Hasan Jony, Rubina Khatun, K.M. Mozaffor Hossain

**Author notes:** Correspondence to: Dr. K.M. Mozaffor Hossain, Professor, Department of Veterinary & Animal Sciences Faculty of Veterinary & Animal Sciences University of Rajshahi, Rajshahi-6205, Bangladesh.

## Abstract

**Background:** Retail red meat may act as a source of foodborne pathogens, antimicrobial-resistant bacteria, and antibiotic residues, posing a significant public health concern in Bangladesh.

**Objectives:** This study aimed to isolate and identify major bacterial pathogens from retail red meat, determine their antimicrobial susceptibility patterns, assess the prevalence of antibiotic-resistant bacteria, and detect antibiotic residues in meat samples.

**Methods:** A cross-sectional study was conducted from January to June 2019 using 60 retail red meat samples (20 cattle, 20 goat, and 20 buffalo) collected from Rajshahi and Naogaon districts. Bacterial isolates were identified using standard cultural, morphological, staining, and biochemical techniques. Antimicrobial susceptibility was evaluated by the Kirby–Bauer disc diffusion method according to CLSI guidelines. Antibiotic residues were screened in 15 representative samples using thin-layer chromatography (TLC).

**Results:** Overall prevalence of Escherichia coli, Salmonella spp., and Staphylococcus aureus was 10.0%, 13.3%, and 28.3%, respectively. E. coli showed complete resistance to penicillin (100%) and high resistance to amoxicillin (83.3%), while remaining highly susceptible to ciprofloxacin (83.3%) and gentamicin (66.7%). Salmonella spp. exhibited highest resistance to penicillin (87.5%) and tetracycline (75.0%), whereas gentamicin (87.5%) and ciprofloxacin (75.0%) remained the most effective agents. S. aureus demonstrated marked resistance to penicillin (94.1%), ampicillin (58.8%), tetracycline (47.1%), and amoxicillin (47.1%), but high susceptibility to gentamicin (88.2%) and ceftriaxone (70.6%). TLC detected ciprofloxacin and oxytetracycline residues in one cattle meat sample each (6.7%).

**Conclusions:** Retail red meat marketed in the study areas harbored multidrug-resistant bacterial pathogens and detectable antibiotic residues, highlighting potential risks to food safety and public health. Continuous surveillance, prudent antimicrobial use, improved slaughterhouse hygiene, and strict compliance with antibiotic withdrawal periods are essential to minimize antimicrobial resistance and residue contamination.

## 1. Introduction

Red meat has been an important part of the human diet throughout human evolution. The term refers to all mammalian muscle meat, including beef, veal, pork, lamb, mutton, horse, buffalo and goat, that is, the flesh, skeletal muscle and any attached connective tissue or fat, excluding bone and bone marrow (Williams, 2007). More broadly, meat refers to animal tissue used as food, mostly skeletal muscle and associated fat, but may also include organs such as lungs, liver, skin, brain, bone marrow, kidney and other internal organs as well as blood (Hammer, 1997). Red meat provides a good source of high-quality protein, beneficial fatty acids and a variety of micronutrients, containing on average 20–24 g of protein per 100 g when raw, and can therefore be considered a high source of protein for the general population, including infants around the time of weaning (Komba et al., 2012).

Beef is a high source of protein containing all the essential amino acids and iron in the heme form, which is better absorbed than the non-heme iron found in plants (Schnepf, 2007). The buffalo carcass has less fat and bone and a higher proportion of muscle than cattle (Sherikar, 2011), while goat meat is increasingly consumed worldwide owing to its distinctive taste and desirable chemical composition. However, meat is easily perishable because it provides a suitable medium for the growth of various micro-organisms (Komba et al., 2012).

The main sources of meat contamination are the slaughtered animals themselves, the workers and the working environment and, to a lesser degree, contamination from air via aerosols and from carcass dressing water (Birhanu et al., 2017). Contaminating organisms are derived mainly from the hide of the animal and comprise organisms originating from the stomach and intestine that are excreted in the faeces (Norrung et al., 2009). Although such pathogens usually cause self-limiting gastroenteritis, invasive disease and complications may also occur; systemic salmonellosis can be life-threatening, and Shiga toxin-producing E. coli, particularly E. coli O157:H7, can cause bloody diarrhoea and haemolytic uraemic syndrome (Griffin, 1995). Intrinsic factors (moisture content, pH, nutritive value, absence of inhibitory substances) and extrinsic factors (temperature, relative humidity, oxygen availability and other chemical and physical properties) affect microbial growth in raw meat and meat products (Adams and Moss, 1999).

Food-borne diseases result from ingestion of bacteria, toxins and cells produced by micro-organisms present in food (Okonko et al., 2010). The most important food-borne bacterial pathogens associated with meat are Salmonella spp., Staphylococcus aureus, Escherichia coli, Campylobacter jejuni, Listeria monocytogenes, Clostridium perfringens, Yersinia enterocolitica and Aeromonas hydrophila (Bhandare et al., 2007), of which Salmonella spp., Campylobacter jejuni, Listeria monocytogenes and verocytotoxin-producing E. coli O157 constitute major public health problems (Griffin, 1995). Lack of knowledge and awareness among traders who handle and distribute meat regarding safety, health, quality and halal requirements can lead to various diseases caused by E. coli and Salmonella spp.

Food safety is one of the critical issues in international trade (Unnevehr, 2000), and many diseases are transmitted through food, sometimes leading to death (Quintavalla and Vicini, 2002). Between 24 and 81 million cases of food-borne disease are reported annually worldwide, of which about 50% are related to animal products (Gravani, 1987). Setting microbiological criteria that indicate an unacceptable food-safety risk is difficult owing to improper sampling and the absence of clear food-safety objectives for pathogens on fresh meat, and public-health awareness of zoonotic food-borne pathogens transmitted from animal-derived food continues to grow (Zhao et al., 2001). Furthermore, high consumption of red and processed meat is linked with an increased risk of colorectal cancer, and adults consuming more than 90 g of red and processed meat daily are advised to reduce intake to an average of 70 g/day cooked weight (NHS Choices, 2011).

The emergence of antibiotic resistance among pathogenic and commensal bacteria has become a serious worldwide problem. The use and overuse of antibiotics in human medicine and animal husbandry create selective pressure that favours the emergence of antimicrobial resistance among micro-organisms (Acar and Moulin, 2006). Although a direct causal role of drug-resistant bacteria in food items in increasing clinical cases of resistant infection is difficult to prove, the presence of such bacteria in food items and their related environment could contribute to the spread of antimicrobial resistance among food-borne pathogens (Farzana et al., 2011), and the prevalence of multidrug-resistant food-borne pathogens increases through consumption of contaminated food, since they cause more serious disease than susceptible bacteria (Gwida and El-Gohary, 2015). Where antibiotic resistance renders treatment ineffective, infection persists and illness progresses (Brunelle et al., 2013).

Judicious antibiotic use, public education on the health risks of promiscuous drug use in livestock production, and hygienic practice at slaughterhouses can help reduce bacterial drug resistance in both man and animals. Antibiotic residues in meat and meat products pose potential health hazards, including allergic skin conditions, nausea, vomiting, anaphylactic shock and even death, and cooking or freezing has minimal effect on such residues. In developing countries such as Bangladesh, antibiotics are frequently used without proper prescription, and much of the public is unaware of the appropriate dosage schedule, often discontinuing a course after one or two days without any awareness of the resulting risk of bacterial resistance. It is therefore essential to isolate and identify bacteria from red meat and to determine the prevailing antibiotic-resistance profile.

Considering the above, the present study was undertaken with the following objectives: (i) to isolate and identify bacteria from red meat; (ii) to determine the antibiotic sensitivity and resistance pattern of the isolated bacteria; (iii) to determine the prevalence of antibiotic-resistant bacteria in red meat; and (iv) to detect antibiotic residues in red meat.

### 1.1 Background and rationale

Meat was incorporated into the human diet at least 2.6 million years ago and played a key evolutionary role, contributing to the development of the unusually large and complex human brain (Pobiner, 2013). Meat is rich not only in protein but in complete and balanced essential amino acids (Schurgers and Vermeer, 2000), and continues to provide high-quality protein, beneficial fatty acids and micronutrients essential for optimal health (Williams, 2007). Red meat is commonly considered to include beef, pork, lamb and game, whereas processed meat is any meat preserved by salting, smoking, curing or the addition of chemical preservatives, such as bacon, sausage, salami or ham (Larsson and Orsini, 2014).

The rate of meat spoilage depends on the initial microbial load (Nursiani, 2003); meat and meat products are frequently contaminated with micro-organisms after leaving the processing plant and during subsequent handling, particularly where hygiene monitoring during non-industrial processing is poor (Stagnitta et al., 2006). Although the meat of healthy animals is essentially sterile, contamination may occur during slaughter, preparation and transport, with the dominant microbial species varying according to pH, oxygen availability, water activity and storage temperature (Ercolin et al., 2006).

Foodborne infection and the resulting illness remain among the major international public-health challenges, contributing to high morbidity, mortality and economic loss even in industrialised countries (Clarence et al., 2009). Microbial contamination reduces the shelf-life of food and promotes food-borne illness; pathogens such as Salmonella spp. and E. coli originating from the animal at slaughter contaminate the carcass and are subsequently spread to cut or raw meat during further processing, creating a major public-health concern (Lecos, 1987). A recent meta-analysis found 18% and 16% higher risk of cardiovascular mortality among the highest categories of processed– and red-meat consumers, respectively, compared with the lowest categories, and a 13% higher risk of type 2 diabetes per 100 g of red meat consumed (Abete et al., 2014). The Global Burden of Disease study attributes approximately 841,000 deaths annually to diets high in processed meat and about 38,000 deaths annually to red meat consumption worldwide (Lim et al., 2013).

The Enterobacteriaceae group, particularly E. coli and Klebsiella sp., are among the most challenging bacterial contaminants of raw and processed meat products worldwide, and their control requires investigation of the causative agents in meat to protect public health (Al-Mutairi, 2011). Raw retail meats are potential vehicles for the transmission of food-borne disease, causing considerable illness, death and economic cost in developing countries (Fratamico et al., 2005). Salmonella sp. accounts for a substantial proportion of food-borne outbreaks; in the United States, Salmonella sp. was reported to account for 48% of all beef-related outbreaks (Bean and Griffin, 1990). Various Salmonella serotypes have been isolated from meat products in different countries, and multiple studies confirm the widespread presence of E. coli, Salmonella sp. and Staphylococcus aureus in raw meat samples worldwide (Mehrabian and Jaberi, 2007; Young et al., 2007).

Staphylococcus aureus, including methicillin-resistant strains (MRSA), can colonise food animals asymptomatically and act as a reservoir or transmission vehicle, with contamination of meat and dairy products reported in several countries (Dinges et al., 2000). Isolation rates of MRSA on retail meat have been reported to vary by meat species, being highest in turkey and lowest in game and fowl (Gill et al., 1998). Salmonella sp. remains among the most important food-borne pathogens worldwide, with outbreaks linked to a wide range of foods including poultry, eggs, beef, fish and dairy products (Izat et al., 1990).

Multiple studies report varying antibiotic-resistance profiles among meat-borne isolates. The highest resistance of E. coli has been reported to amoxicillin–clavulanic acid, norfloxacin, cephalothin and nalidixic acid, with comparatively low resistance to ertapenem, aztreonam and gentamicin (Abdellah et al., 2013). Reported S. aureus prevalence and multidrug-resistance patterns from various food sources commonly implicate cephalosporins, fluoroquinolones, beta-lactams and aminoglycosides (Hemalata and Virupakshaiah, 2016), while several antibiotic classes, including tetracyclines, sulfonamides, fluoroquinolones, macrolides, lincosamides, aminoglycosides, beta-lactams and cephalosporins, are extensively administered to food-producing animals (Jank et al., 2017).

Antibiotic residues in meat also constitute a distinct public-health hazard. Reported prevalence of antibiotic residues in meat has varied considerably across countries, from 7.4% in Vietnam to 57.7% in Turkey (Mensah et al., 2014). Possible adverse effects of residues include allergic and anaphylactic reactions, chronic toxicity from prolonged low-level exposure, development of antibiotic-resistant bacteria in treated animals, and disruption of normal human intestinal flora (Hamann et al., 1979). Tetracycline residues in the human diet have been linked with poor foetal development, staining of teeth in young children, gastrointestinal disorders and pro-inflammatory, cytotoxic and immuno-pathological effects (Lawal et al., 2015), while residues of sulphamethazine, oxytetracycline and furazolidone have been associated with immuno-pathological effects, and gentamicin and chloramphenicol residues with mutagenic, nephropathic and hepatotoxic effects (Nisha, 2008). Continuous exposure to ciprofloxacin residues has also been linked with toxicity and impaired cytochrome (CYP1A2)-mediated drug metabolism (Khan et al., 2015).

## 2. Materials and Methods

The research was conducted in the Microbiology Laboratory, Department of Veterinary and Animal Sciences, University of Rajshahi, Bangladesh, during the period January to June 2019.

### 2.1 Study area and sample collection

The study was conducted in randomly selected retail meat shops in four upazila of Rajshahi district (Charghat, Durgapur, Godagari and Paba) and the Rajshahi City Corporation area, and in five upazila of Naogaon district (Badalgachi, Manda, Mohadevpur, Patnitala and Naogaon Sadar). A total of 60 red meat samples were collected: 30 from Rajshahi district and 30 from Naogaon district. From each of the ten study areas, two cattle-meat, two goat-meat and two buffalo-meat samples were collected, giving a total of 20 cattle-meat, 20 goat-meat and 20 buffalo-meat samples overall (Table 1).

**Table 1.**
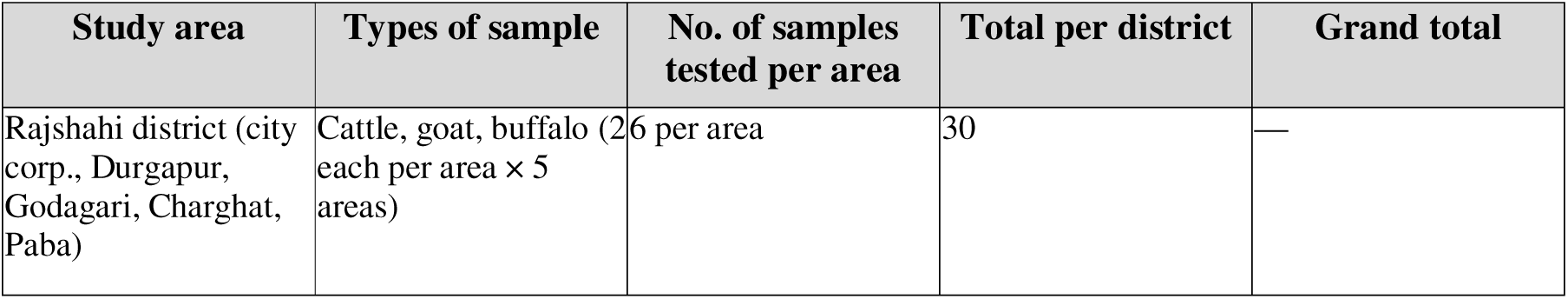
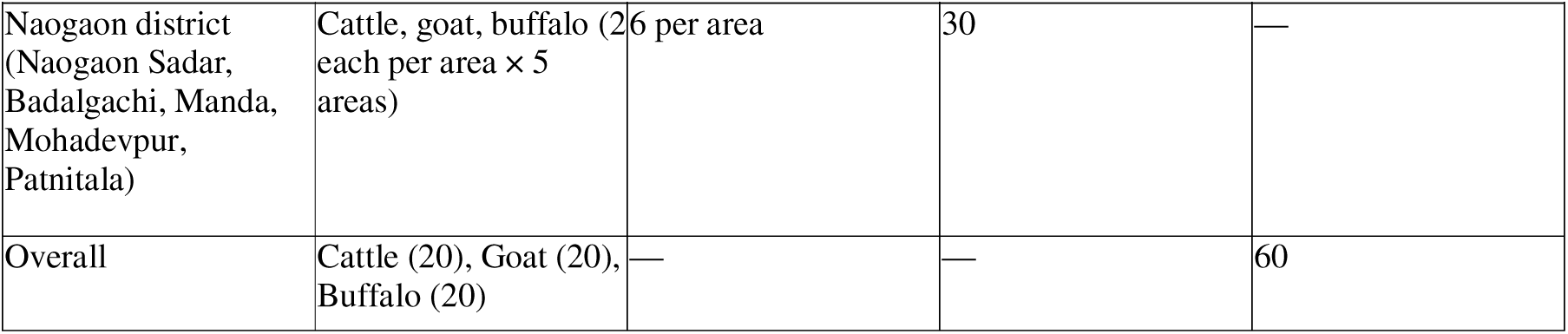
Basic information of study samples.

Samples were collected aseptically from retail meat shops, with precautions taken to prevent cross-contamination. Each sample was placed in a sterile zip-lock bag with an identification mark and transported to the laboratory in an ice-containing thermo flask, where it was examined as soon as possible.

### 2.2 Media, chemicals and antibiotic discs

Liquid media used included Nutrient broth, Methyl-Red and Voges-Proskauer (MR-VP) broth, peptone water broth and sugar media. Solid media used included Nutrient agar, Blood agar, MacConkey agar, Eosin Methylene Blue (EMB) agar, Triple Sugar Iron (TSI) agar, Salmonella-Shigella (SS) agar, Mannitol Salt Agar (MSA) and Mueller-Hinton agar (MHA), all prepared according to the manufacturers’ instructions and sterilised by autoclaving at 121°C and 15 psi for 15 minutes. Gram’s stain reagents, methyl red and Voges-Proskauer solutions, Kovac’s reagent, phosphate-buffered saline (PBS), 50% sterile buffered glycerine and 3% hydrogen peroxide were also prepared. Thirteen commercially available antibiotic discs (Difco, USA) were used, of which seven, commonly used in the field for treatment of animals and birds, were selected for the antibiogram assay: penicillin (10 µg), gentamicin (10 µg), tetracycline (30 µg), ampicillin (25 µg), amoxycillin (30 µg), ciprofloxacin (5 µg) and ceftriaxone (30 µg).

**Figure 1.**
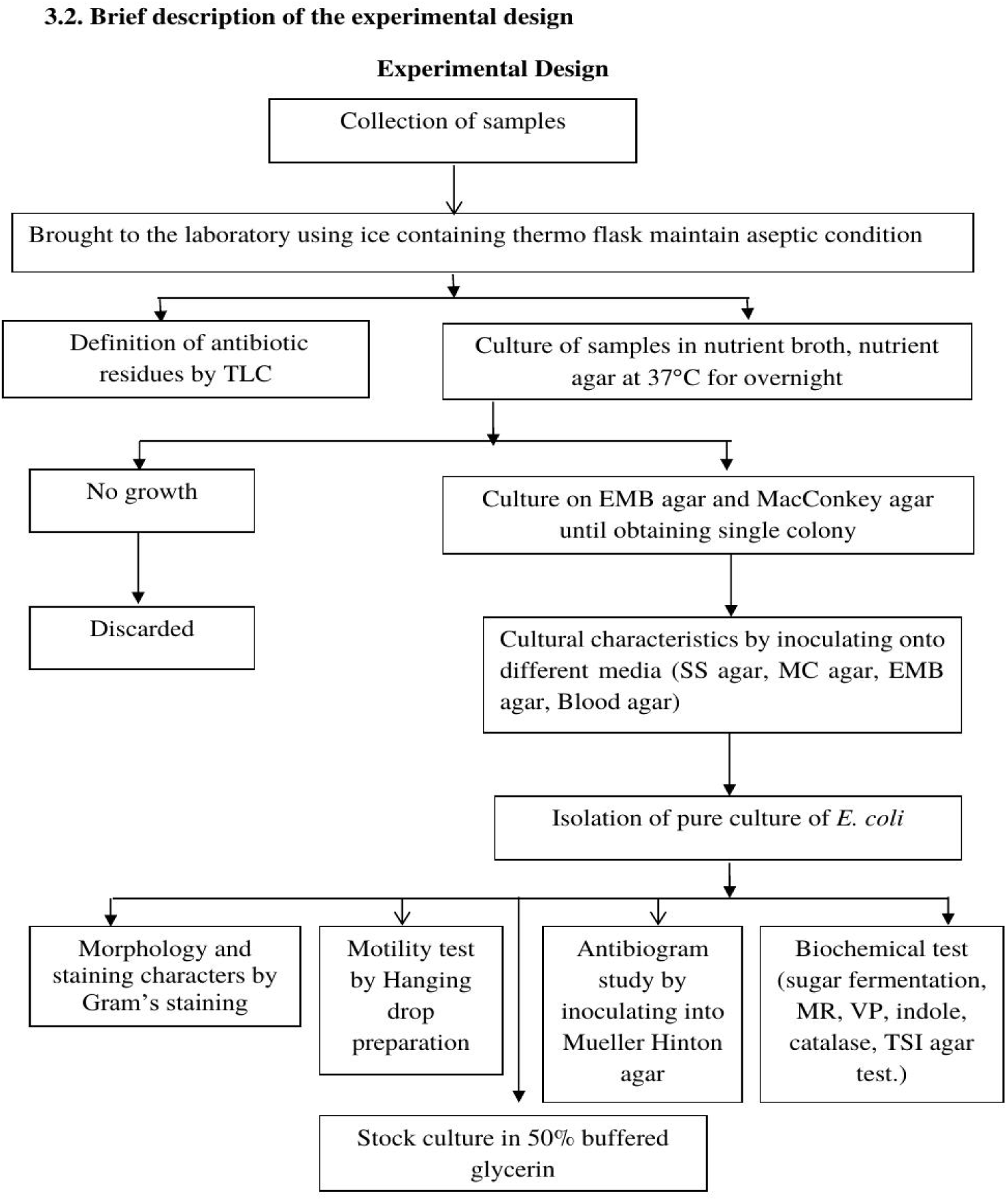
Flow-chart showing the design of the experiment.

### 2.3 Sample preparation, isolation and identification of bacteria

Swabs applied to meat samples were submerged in sterile saline as a ‘meat wash’, which was used to inoculate appropriate enrichment broth. After enrichment in nutrient broth, a loopful of culture was streaked onto EMB, SS and MSA and incubated aerobically at 37°C for 24 hours. Presumptive colonies were sub-cultured to obtain pure isolates. Colony morphology (shape, size, surface texture, edge, elevation, colour and opacity) was recorded following Merchant and Packer (1967). Gram’s staining was performed according to Cheesbrough (1981) and examined under a compound microscope at 100× magnification using immersion oil. Motility was assessed by the hanging-drop method described by Cowan (1965).

### 2.4 Biochemical characterisation

Pure isolates were subjected to a battery of biochemical tests: catalase test (3% H2O2, observed for bubble formation), methyl red (MR) test, Voges-Proskauer (VP) test, indole test (Kovac’s reagent), sugar fermentation with five basic sugars (dextrose, lactose, sucrose, maltose and mannitol) in peptone water containing phenol red indicator and inverted Durham’s tubes, and Triple Sugar Iron (TSI) agar slant reaction, all performed according to standard procedures (Cheesbrough, 1981).

### 2.5 Antibiotic susceptibility testing

Antimicrobial susceptibility of the isolates was determined by the Kirby-Bauer disc diffusion method (Bauer et al., 1966) on Mueller-Hinton agar, following the recommendations of the Clinical and Laboratory Standards Institute (CLSI, 2016). A standardised broth culture (incubated overnight at 37°C) was spread evenly onto MHA plates, and antimicrobial discs were applied at approximately 1 cm apart. Plates were incubated at 37°C for 16–18 hours, and zones of inhibition were measured to the nearest millimetre and interpreted as resistant (R), intermediate (I) or sensitive (S) according to CLSI (2016) breakpoints (Table 2).

**Table 2.** Zone-diameter interpretative standards for antimicrobial susceptibility.

| Antibiotic | Symbol | Disc concentration (µg/disc) | Resistant (mm) | Intermediate (mm) | Sensitive (mm) |
| --- | --- | --- | --- | --- | --- |
| Penicillin | P | 10 | ≤11 | 12–21 | ≥22 |
| Gentamicin | GEN | 10 | ≤12 | 13–14 | ≥15 |
| Tetracycline | TE | 30 | ≤11 | 12–14 | ≥15 |
| Ampicillin | AMP | 25 | ≤13 | 14–16 | ≥17 |
| Amoxycillin | AMX | 30 | ≤13 | 14–17 | ≥18 |
| Ciprofloxacin | CIP | 5 | ≤15 | 16–20 | ≥21 |
| Ceftriaxone | CRO | 30 | ≤13 | 14–20 | ≥21 |

Pure isolates were preserved in 50% sterile buffered glycerine at −20°C for maintenance of stock cultures.

### 2.6 Detection of antibiotic residues by thin-layer chromatography

Antibiotic residues in meat samples were screened by thin-layer chromatography (TLC). A volume of 20 µL of standard solution and sample extract was spotted onto silica TLC plates using an automatic TLC spotter. Chromatographic chambers were saturated with the mobile phase (acetone/methanol, 1:1) for 30 minutes before development. After development, plates were dried and examined under short-wave (254 nm) and long-wave (365 nm) ultraviolet light, and fluorescing or absorbing spots were marked and compared with reference standards.

## 3. Results

### 3.1 Cultural, morphological and staining characteristics

On nutrient broth, growth of all three organisms was indicated by diffuse turbidity, with pellicle formation occasionally observed. On nutrient agar, E. coli produced smooth, circular, white-to-greyish-white colonies; Salmonella sp. produced smooth, low convex, greyish-white translucent colonies; and S. aureus produced smooth, large, circular, shiny, golden-yellow colonies. On EMB agar, E. coli produced greenish-black colonies with a characteristic metallic sheen, while Salmonella sp. produced translucent, amber-coloured colonies; S. aureus was inhibited on EMB agar. On MacConkey agar, E. coli produced bright pink colonies, whereas Salmonella sp. produced transparent, colourless colonies with no zone of bile-salt precipitation; S. aureus was again inhibited. On Salmonella-Shigella agar, E. coli produced pink-to-rose-red colonies, while Salmonella sp. produced colourless colonies with a black centre; S. aureus was inhibited. On Mannitol Salt Agar, only S. aureus grew, producing yellow colonies surrounded by yellow zones, while both Gram-negative organisms were inhibited. On blood agar, E. coli produced non-haemolytic colourless colonies, Salmonella sp. produced non-haemolytic smooth white colonies, and S. aureus produced haemolytic, white-to-cream coloured colonies (Table 3, Table 4, Table 5).

**Table 3.** Morphology and cultural characteristics of isolated E. coli.

| Nutrient agar | MacConkey agar | EMB agar | SS agar | Blood agar | Mannitol salt agar | Gram stain / motility |
| --- | --- | --- | --- | --- | --- | --- |
| Circular, smooth, low convex, glistening colonies | Bright pink, transparent, smooth, raised colonies | Greenish-black colony with metallic sheen | Slight pink, smooth colonies | Colourless, non-haemolytic | No growth | Gram-negative, pink rods, single/paired; motile (+) |

**Table 4.**
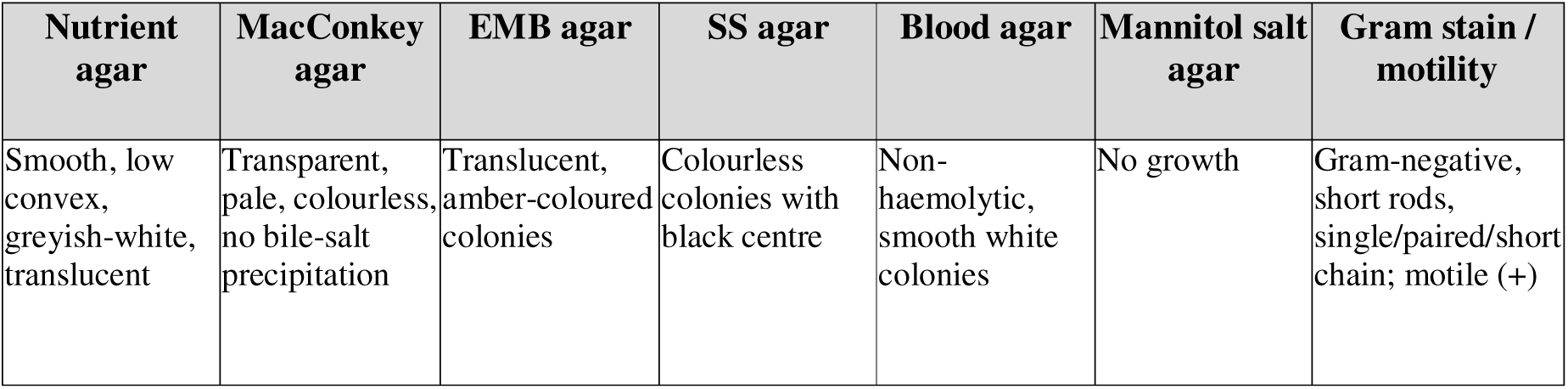
Morphology and cultural characteristics of isolated Salmonella sp.

| Nutrient agar | MacConkey agar | EMB agar | SS agar | Blood agar | Mannitol salt agar | Gram stain / motility |
| --- | --- | --- | --- | --- | --- | --- |
| Smooth, low convex, greyish-white, translucent | Transparent, pale, colourless, no bile-salt precipitation | Translucent, amber-coloured colonies | Colourless colonies with black centre | Non-haemolytic, smooth white colonies | No growth | Gram-negative, short rods, single/paired/short chain; motile (+) |

**Table 5.** Morphology and cultural characteristics of isolated Staphylococcus aureus.

| Nutrient agar | MacConkey agar | EMB agar | SS agar | Blood agar | Mannitol salt agar | Gram stain / motility |
| --- | --- | --- | --- | --- | --- | --- |
| Smooth, large, circular, shiny, golden-yellow | No growth | No growth | No growth | Haemolytic, white/cream colonies | Yellow colonies surrounded by yellow zones | Gram-positive, cocci in grape-like clusters; non-motile (-) |

Gram-stained smears revealed E. coli as Gram-negative, pink-coloured, small rods arranged singly or in pairs; Salmonella sp. as Gram-negative, short rods, single, paired or in short chains; and S. aureus as Gram-positive cocci arranged in grape-like clusters. In the hanging-drop motility test, E. coli and Salmonella sp. were motile, whereas S. aureus was non-motile.

**Figure 2 & 3.** Growth of bacteria in nutrient broth (diffuse cloudiness and heavy sediment; right tube is control) and on nutrient agar (E. coli: smooth, circular, white-to-greyish-white colonies; Salmonella sp.: smooth, low convex, greyish-white translucent colonies; S. aureus: smooth, large, circular, shiny, golden-yellow colonies).

**Figure 2.**
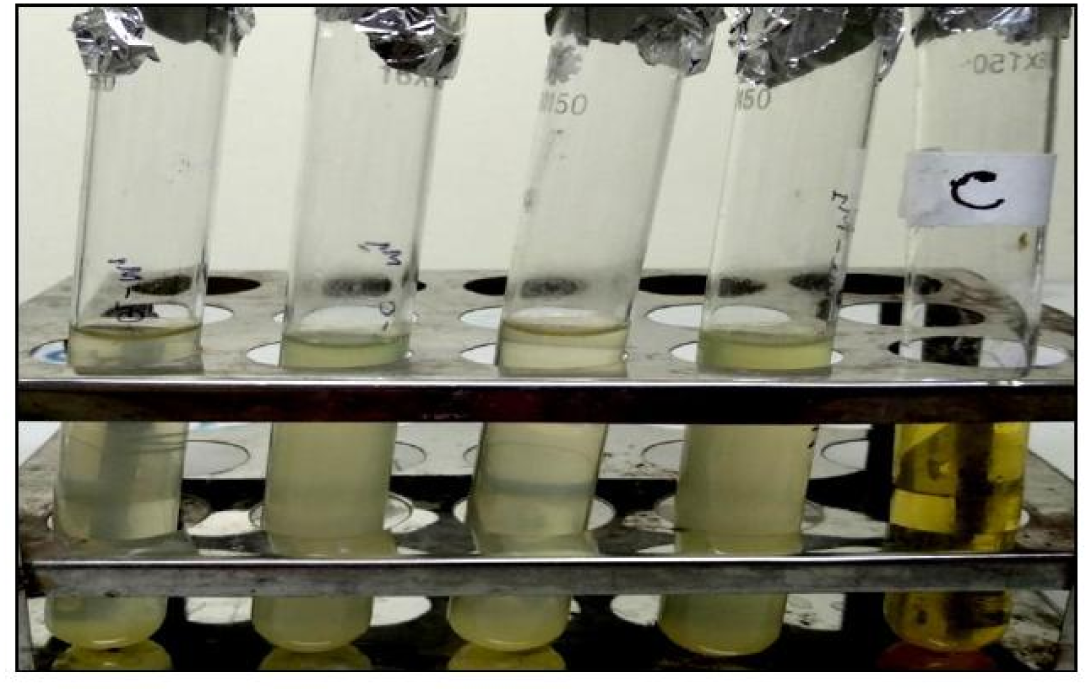
Growth of bacteria in nutrient broth was confirmed by diffuse cloudiness and heavy sediment (right one is control).

**Figure 3.**
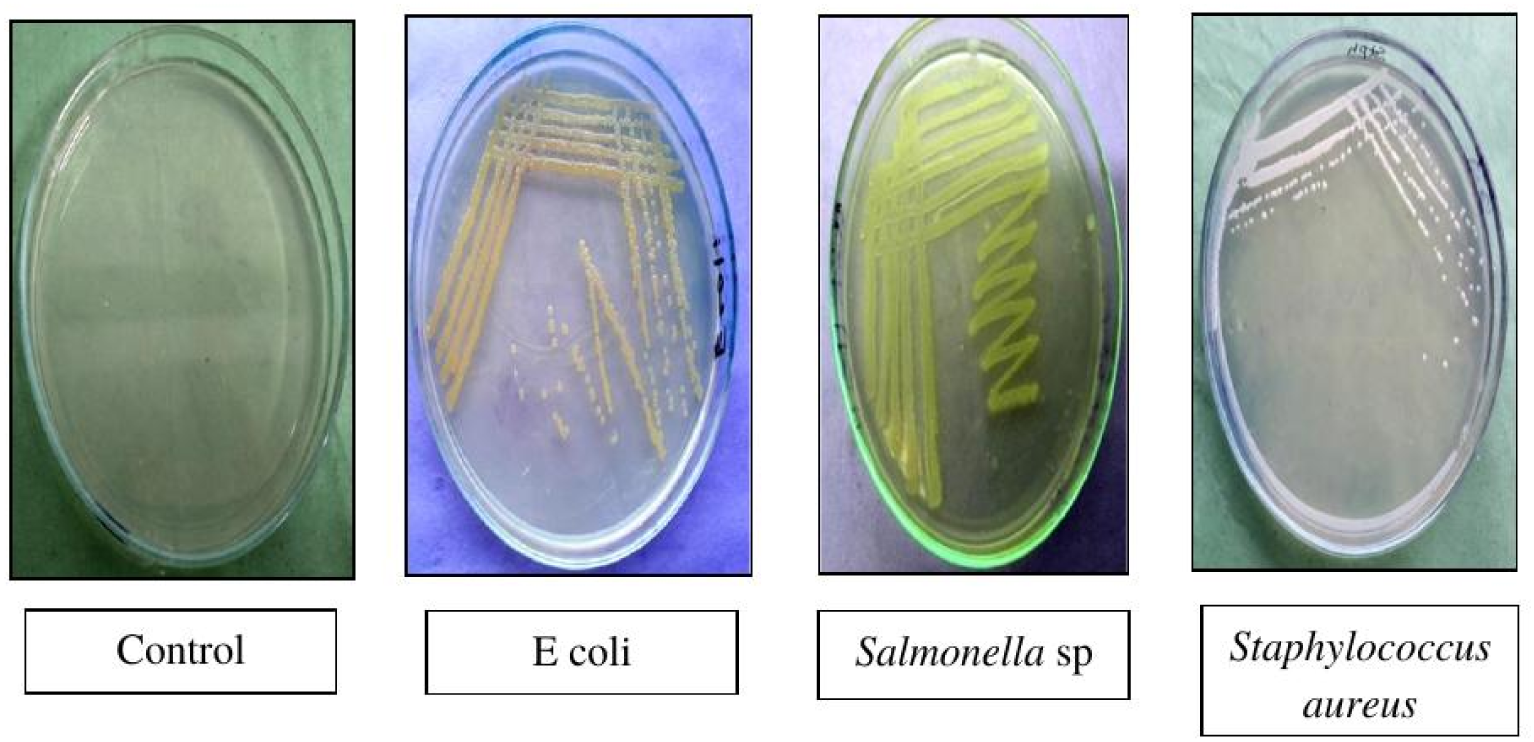
Growth of *E. coli* on nutrient agar indicated by the development of smooth, circular, white to grayish white colony. Growth of *Salmonella* sp by the development of smooth, low convex, grayish white, translucent colony and growth of *Staphylococcus* aureus was indicated by the development of smooth, large circular shiny surface, golden yellow colony (left one is control).

**Figure 4 & 5.** Growth on EMB agar (E. coli: greenish-black colonies with metallic sheen; Salmonella sp.: translucent, amber-coloured colonies) and on MacConkey agar (E. coli: bright pink colonies; Salmonella sp.: transparent, colourless colonies).

**Figure 4.**
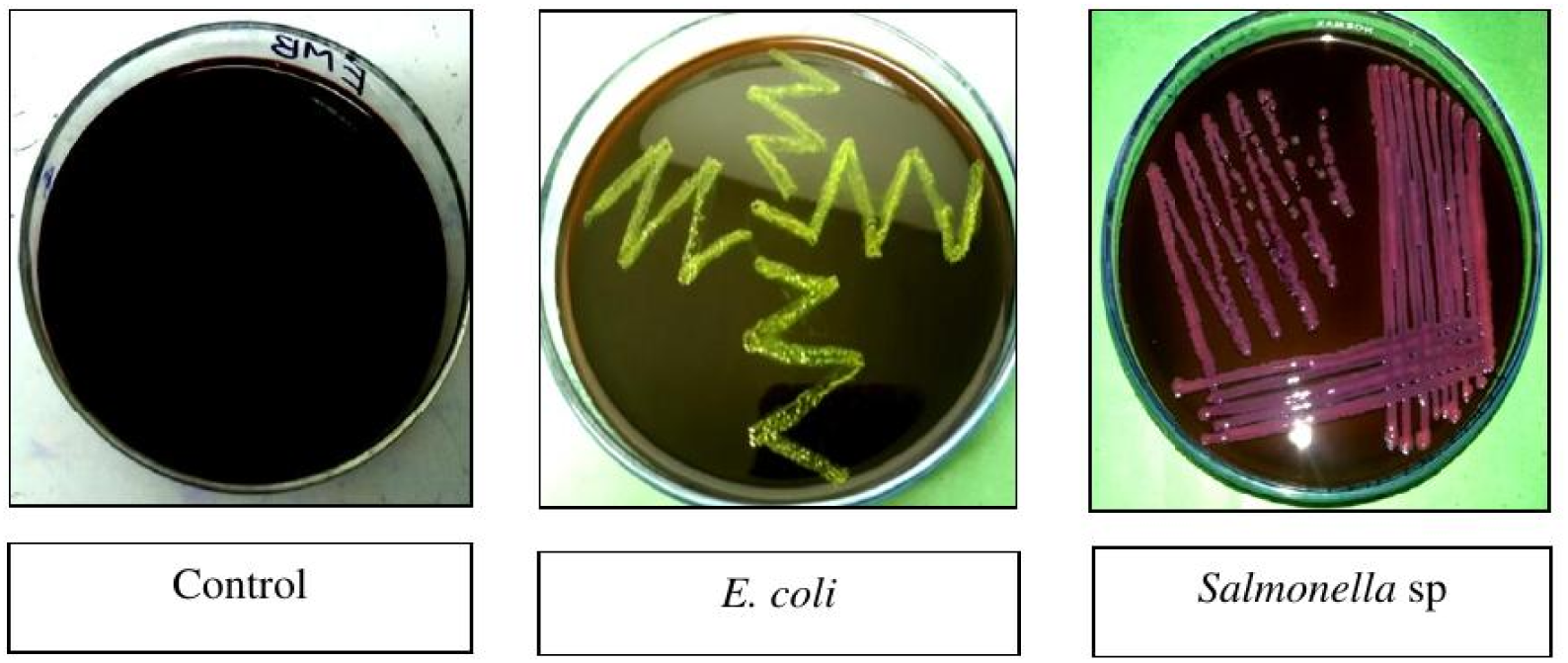
*E. coli* isolates produced greenish-black colonies with metallic sheen on EMB agar and *Salmonella* sp isolates produced translucent, amber colored colonies on EMB agar (left one is control).

**Figure 5.**
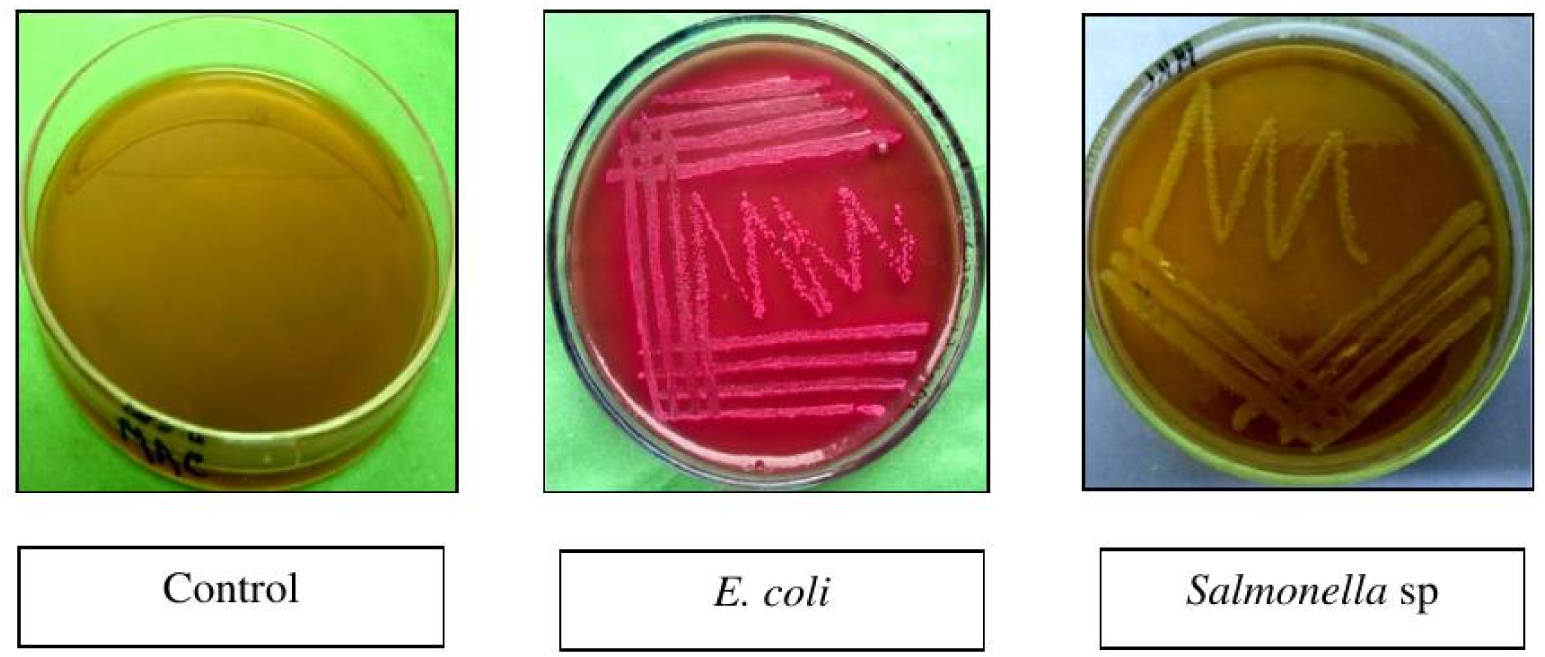
*E. coli* isolates produced smooth, bright pick colored colonies on MacConkey agar and *Salmonella* sp isolates produced transparent, pale and colorless colonies on MacConkey agar (left one is control).

**Figure 6 & 7.** Growth on Salmonella-Shigella agar (E. coli: pink-to-rose-red colonies; Salmonella sp.: colourless colonies with black centre) and on Mannitol Salt Agar (S. aureus: yellow colonies surrounded by yellow zones).

**Figure 6.**
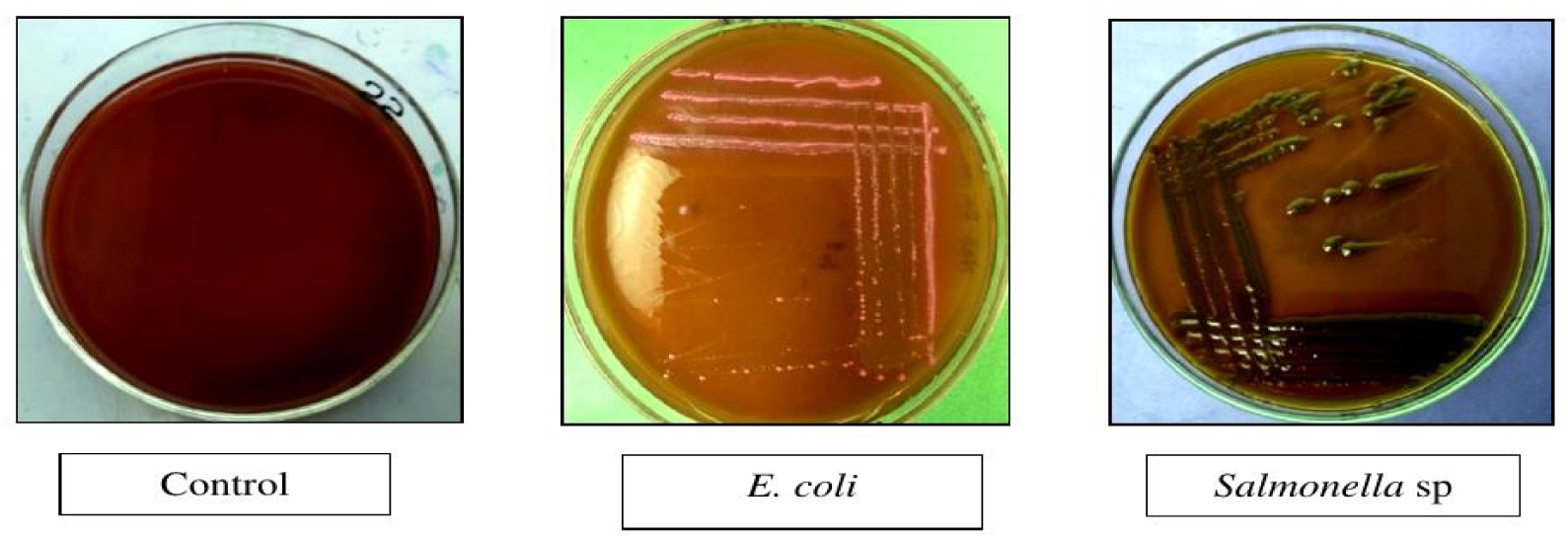
*E. coli* isolates produced circular growth and pink to rose-red colonies on SS agar and *Salmonella* sp isolates produced colorless colonies with black center on SS agar (left one is control).

**Figure 7.**
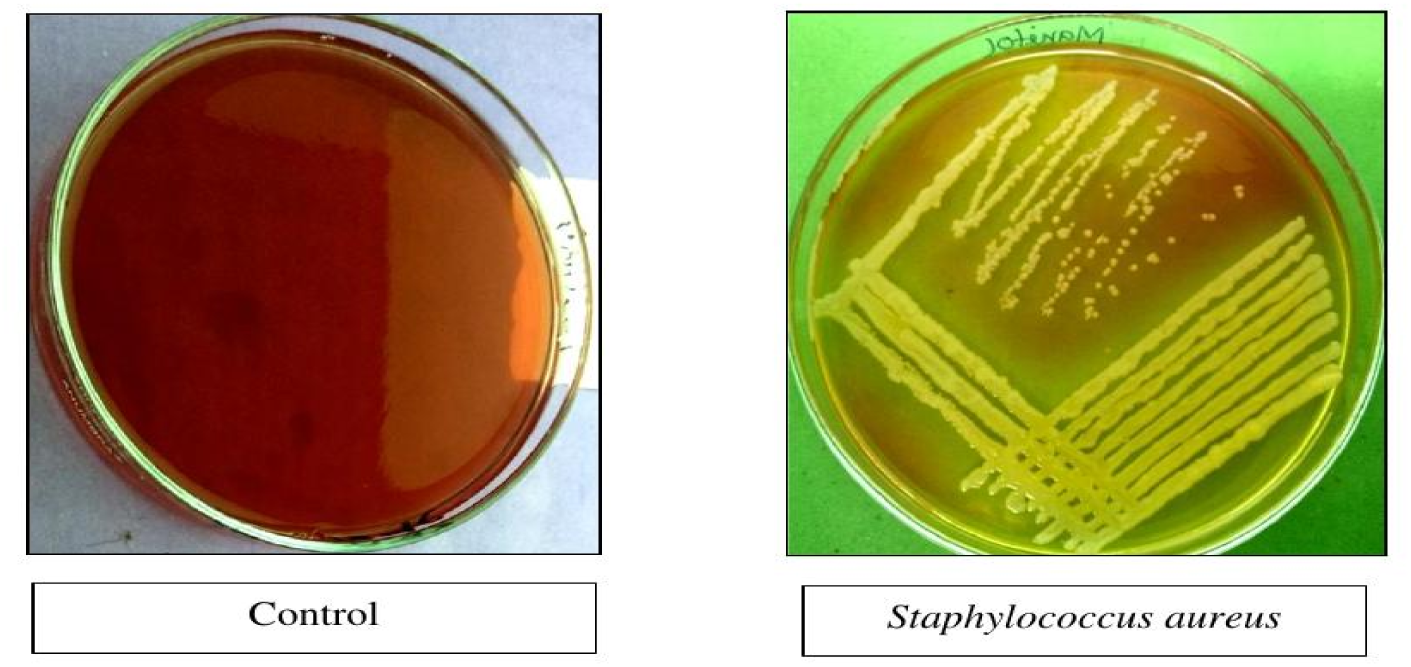
*Staphylococcus aureus* isolates produced yellow colonies surround by yellow zones on MSA agar (left one is control).

**Figure 8 & 9.** Growth on blood agar (E. coli: non-haemolytic colonies; Salmonella sp.: non-haemolytic smooth white colonies; S. aureus: haemolytic white/cream colonies) and Gram-stained micrographs (100×) of E. coli, Salmonella sp. and S. aureus.

**Figure 8.**
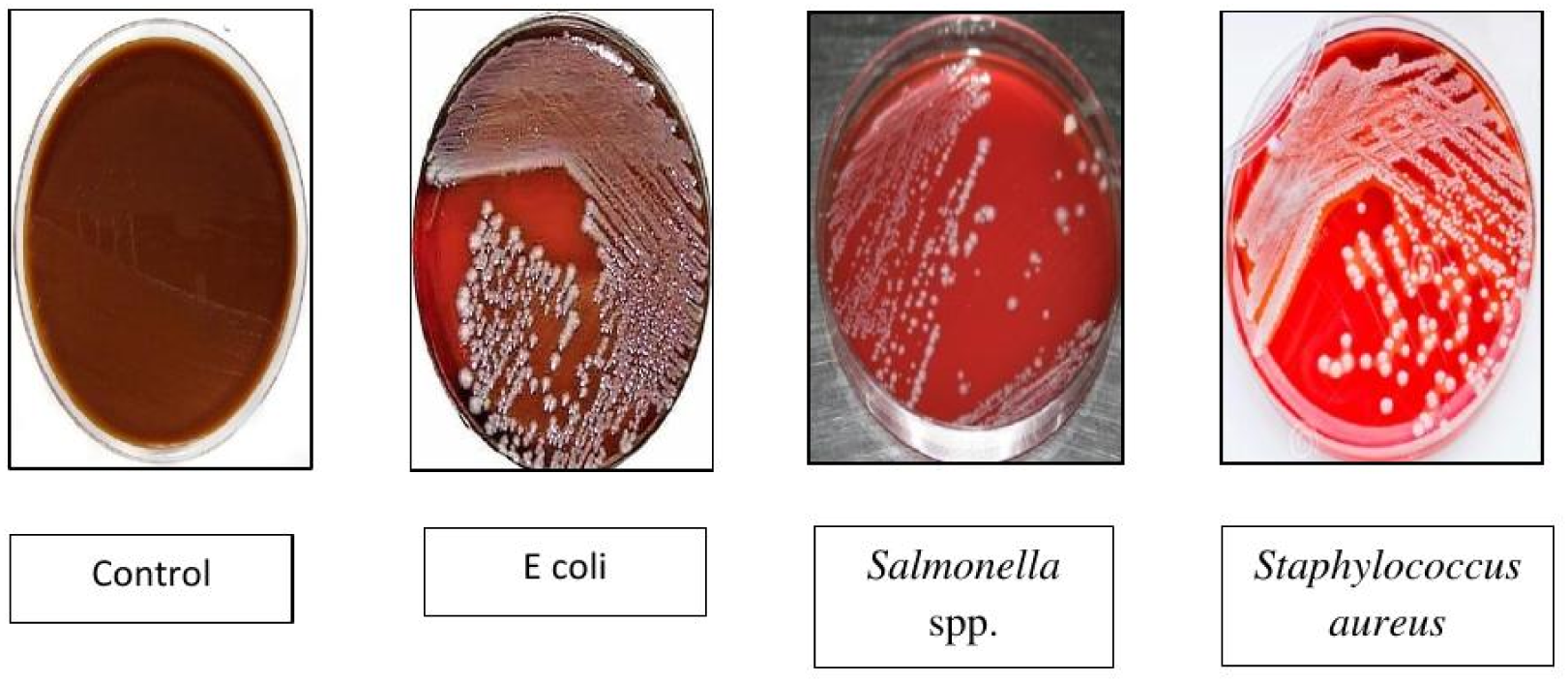
*E. coli* isolates produced non hemolytic colonies on Blood agar, *Salmonella* sp isolates produced non-hemolytic smooth white colonies on blood agar a and *Staphylococcus aureus* isolates produced hemolytic white or cream color colonies ob blood agar (left one is control).

**Figure 9.**
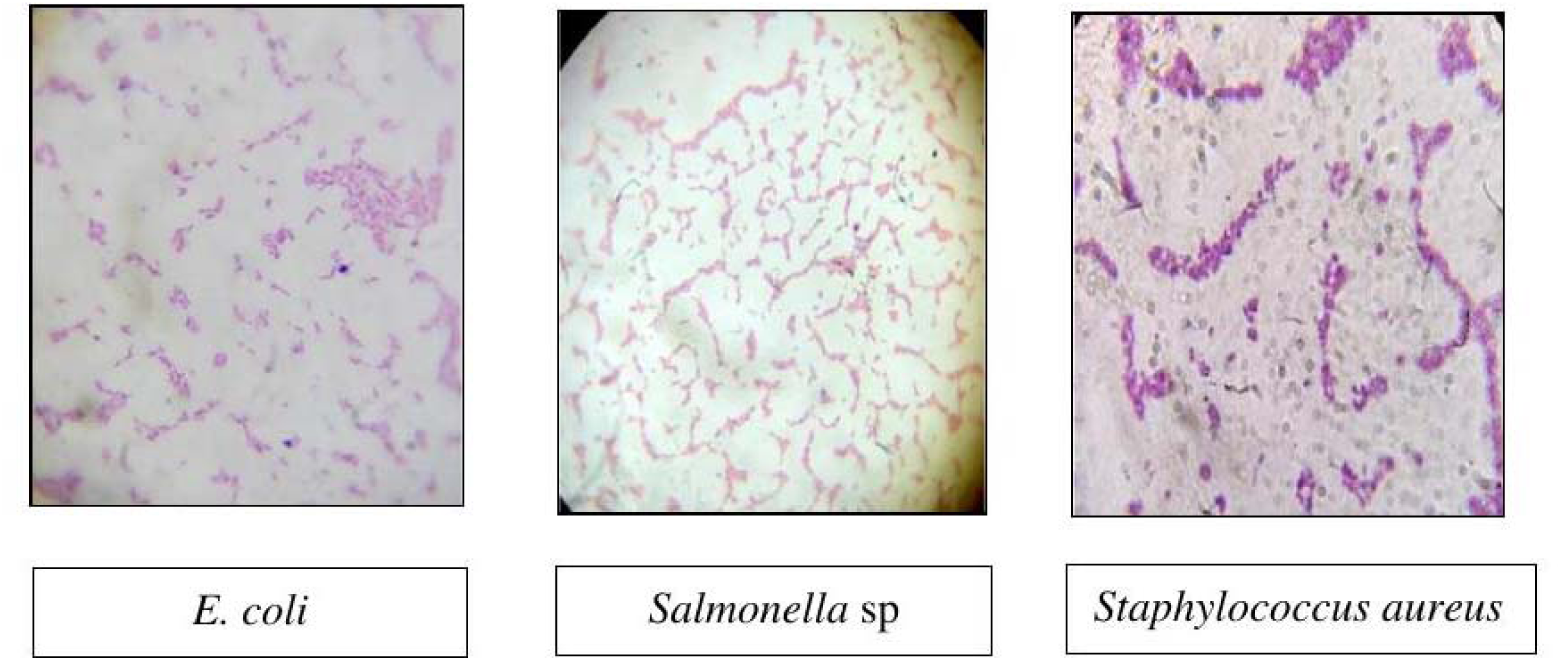
*E. coli* appears as Gram negative, pink colored, rod shaped under a light microscope (100X). *Salmonella* sp appears as Gram negative, short rod-shaped, single, paired or in short chain under a light microscope (100X) and *Staphylococcus aureus* appears as Gram-positive, cocci-shaped and tend to be arranged in clusters that arc described as “grape-like” under a light microscope.

### 3.2 Biochemical characteristics

E. coli fermented dextrose, lactose, sucrose, maltose and mannitol with production of both acid and gas, gave positive indole, catalase and methyl-red reactions, negative Voges-Proskauer and citrate-utilisation reactions, and produced an acidic slant / acidic butt with gas on TSI agar. Salmonella sp. fermented dextrose and mannitol with acid and gas but not lactose or sucrose, gave positive catalase, methyl-red and citrate-utilisation reactions, negative indole and Voges-Proskauer reactions, and produced an alkaline slant / acidic butt with gas (and H2S production) on TSI agar. S. aureus fermented all five sugars with acid but no gas, gave positive catalase, methyl-red, Voges-Proskauer, citrate-utilisation and coagulase reactions, negative indole, and produced an acidic slant / acidic butt with no gas on TSI agar (Table 6, Table 7, Table 8).

**Table 6.** Biochemical properties of isolated E. coli.

| Test | Result |
| --- | --- |
| Sugar fermentation (dextrose, maltose, lactose, sucrose, mannitol) | Acid + gas for all five sugars |
| Indole test | Positive |
| Catalase test | Positive |
| MR test | Positive |
| VP test | Negative |
| TSI slant | Acidic slant / acidic butt with gas |
| Citrate utilisation | Negative |
| Coagulase test | Negative |

**Table 7.** Biochemical properties of isolated Salmonella sp.

| Test | Result |
| --- | --- |
| Sugar fermentation | Dextrose & mannitol: acid + gas; Lactose & sucrose: negative |
| Indole test | Negative |
| Catalase test | Positive |
| MR test | Positive |
| VP test | Negative |
| TSI slant | Alkaline slant / acidic butt with gas (H<sub>2</sub>S+) |
| Citrate utilisation | Positive (royal blue) |
| Coagulase test | Negative |

**Table 8.**
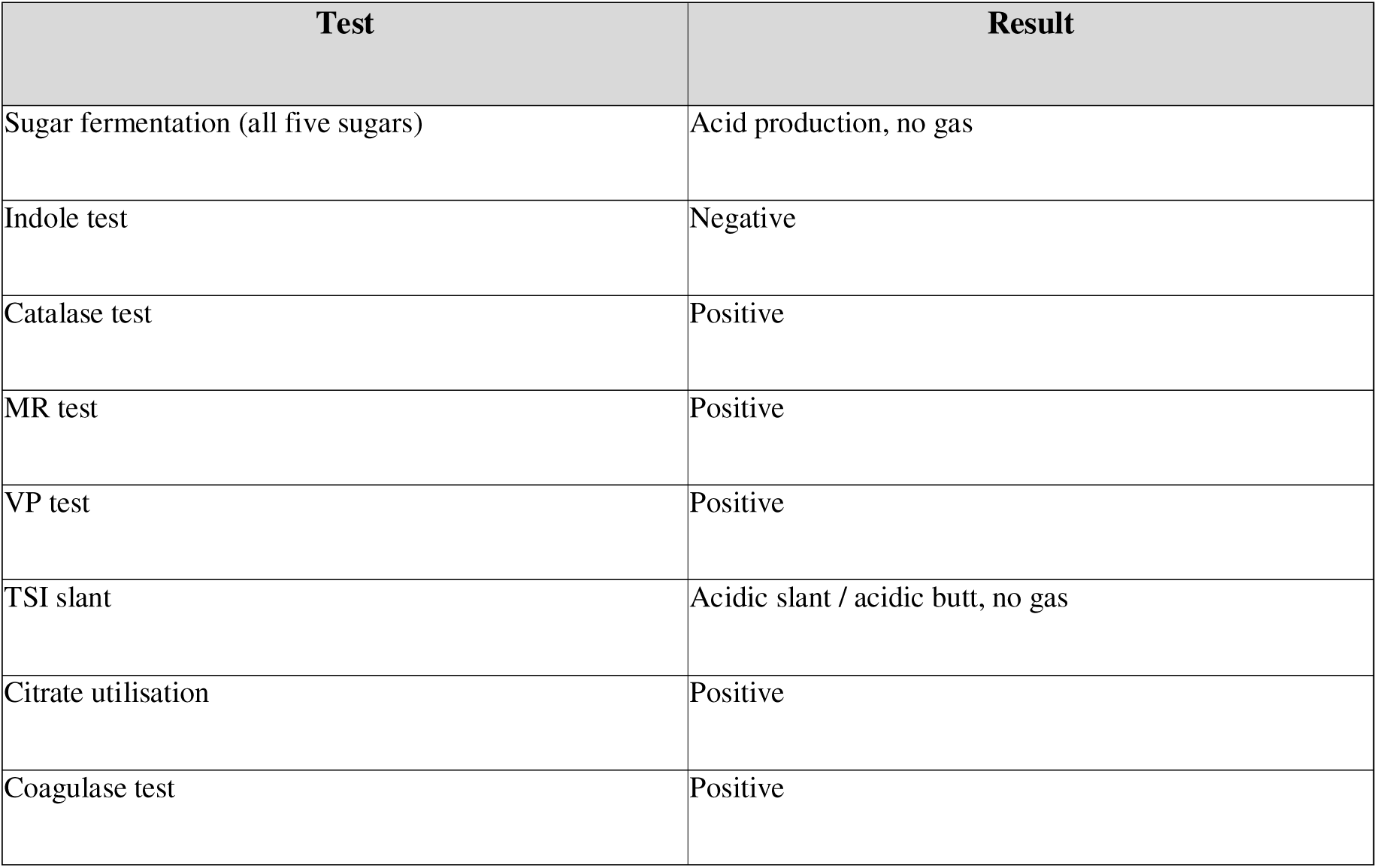
Biochemical properties of isolated Staphylococcus aureus.

| Test | Result |
| --- | --- |
| Sugar fermentation (all five sugars) | Acid production, no gas |
| Indole test | Negative |
| Catalase test | Positive |
| MR test | Positive |
| VP test | Positive |
| TSI slant | Acidic slant / acidic butt, no gas |
| Citrate utilisation | Positive |
| Coagulase test | Positive |

**Figure 10, 11 & 12.** Fermentation of the five basic sugars (dextrose, maltose, sucrose, mannitol, lactose) by E. coli (acid and gas from all five sugars), Salmonella sp. (acid and gas from all except sucrose and lactose) and S. aureus (acid production without gas from all five sugars).

**Figure 10.**
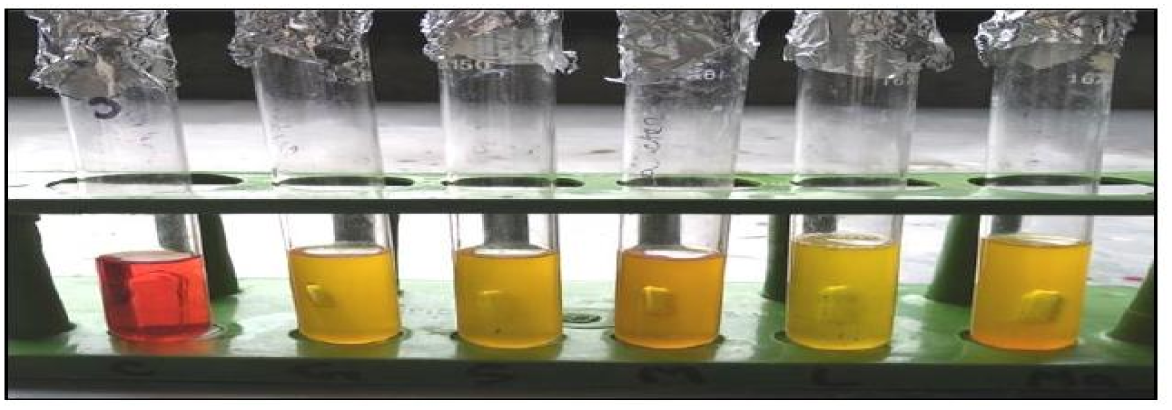
Fermentation activity of *E. coli* with five basic sugars. *E. coli* isolates fermented dextrose, lactose, sucrose, maltose and mannitol with the production of acid and gas (left one is control).

**Figure 11.**
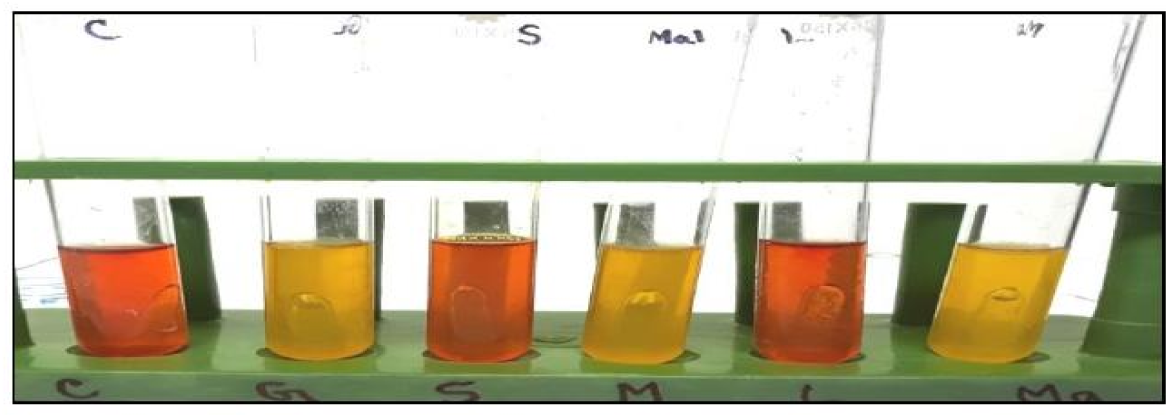
Fermentation activity of *Salmonella* sp with five basic sugars. *Salmonella* sp isolates fermented dextrose, maltose and mannitol with the production of acid and gas except sucrose and lactose (left one is control).

**Figure 12.**
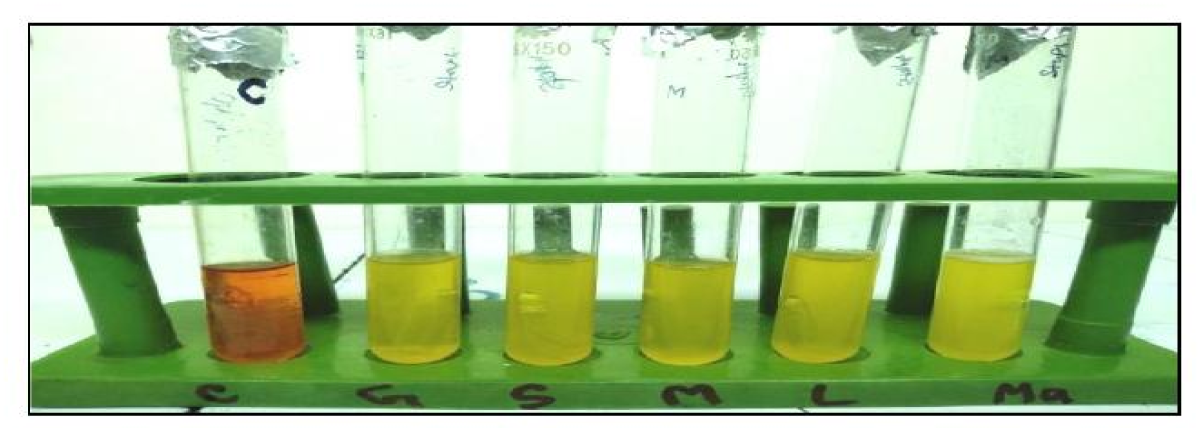
Fermentation activity of *Staphylococcus* aureus with five basic sugars. *Staphylococcus* aureus isolates fermented dextrose, lactose, sucrose, maltose and mannitol with production of acid but no gas (left one is control).

**Figure 13, 14, 15 & 16 (part).** Catalase test (bubble formation, positive), indole test (red/red-violet surface layer in E. coli; negative in Salmonella sp. and S. aureus) and methyl-red (MR) test (red colour development in E. coli and Salmonella sp.).

**Figure 13.**
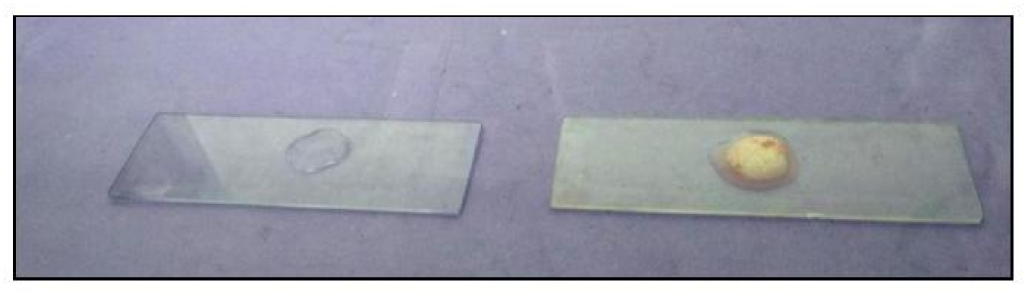
Isolated bacteria showed positive result in Catalase test indicated by production of bubble (left one is control).

**Figure 14.**
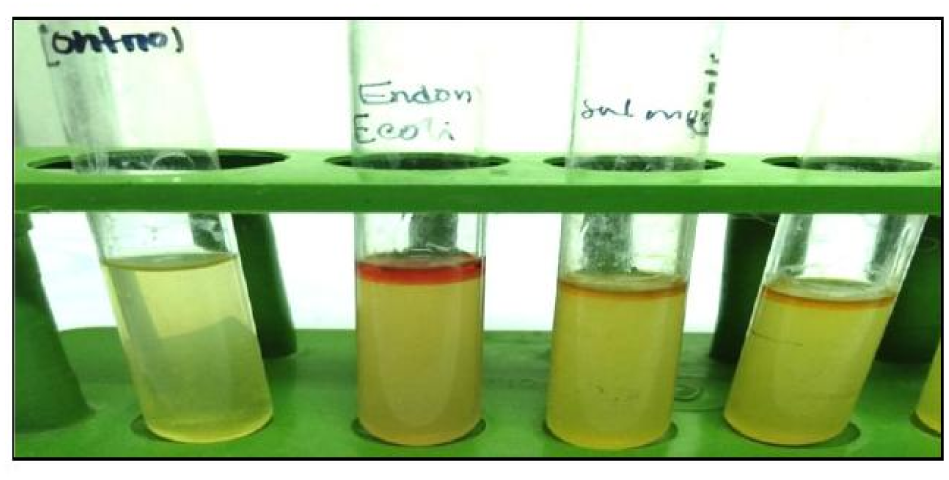
*E. coli* showed positive reaction indicated by development of red or red-violet color in the surface of the alchohol layer if the broth and *Salmonella* and *Staphylococcus* aureus showed negative reaction in Indole test. (left one is control).

**Figure 15.**
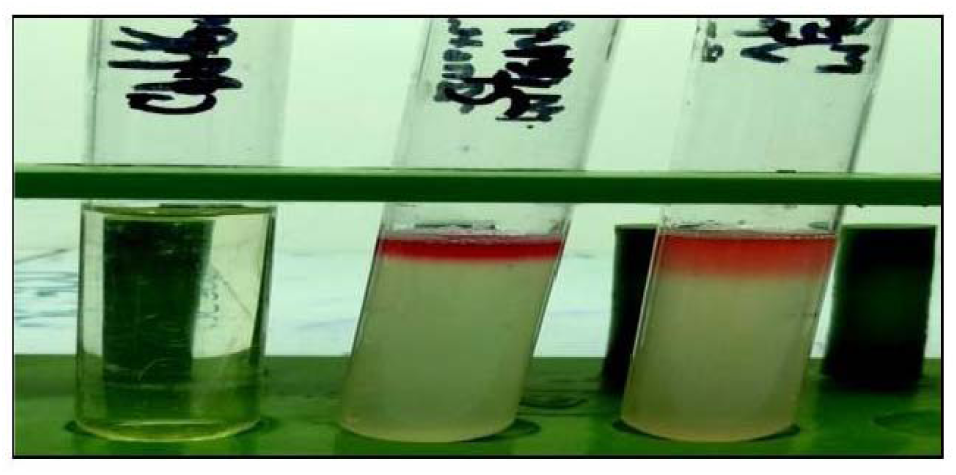
Isolated *E. coli* and *Salmonella* sp showed positive result indicated by the development of red color in MR test (left one is control).

**Figure 16 & 17.** Voges-Proskauer (VP) test (negative in E. coli and Salmonella sp.; positive, red colour, in S. aureus) and Triple Sugar Iron (TSI) agar slant reaction (all three organisms positive).

**Figure 16.**
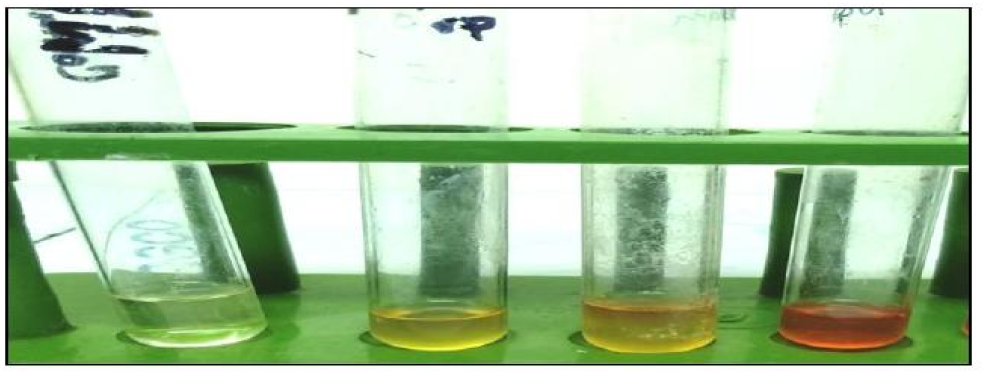
Isolated *E. coli* and *Salmonella* sp showed negative result and Staphylococcus aureus showed positive result indicated by development of red color in VP test (left one is control).

**Figure 17.**
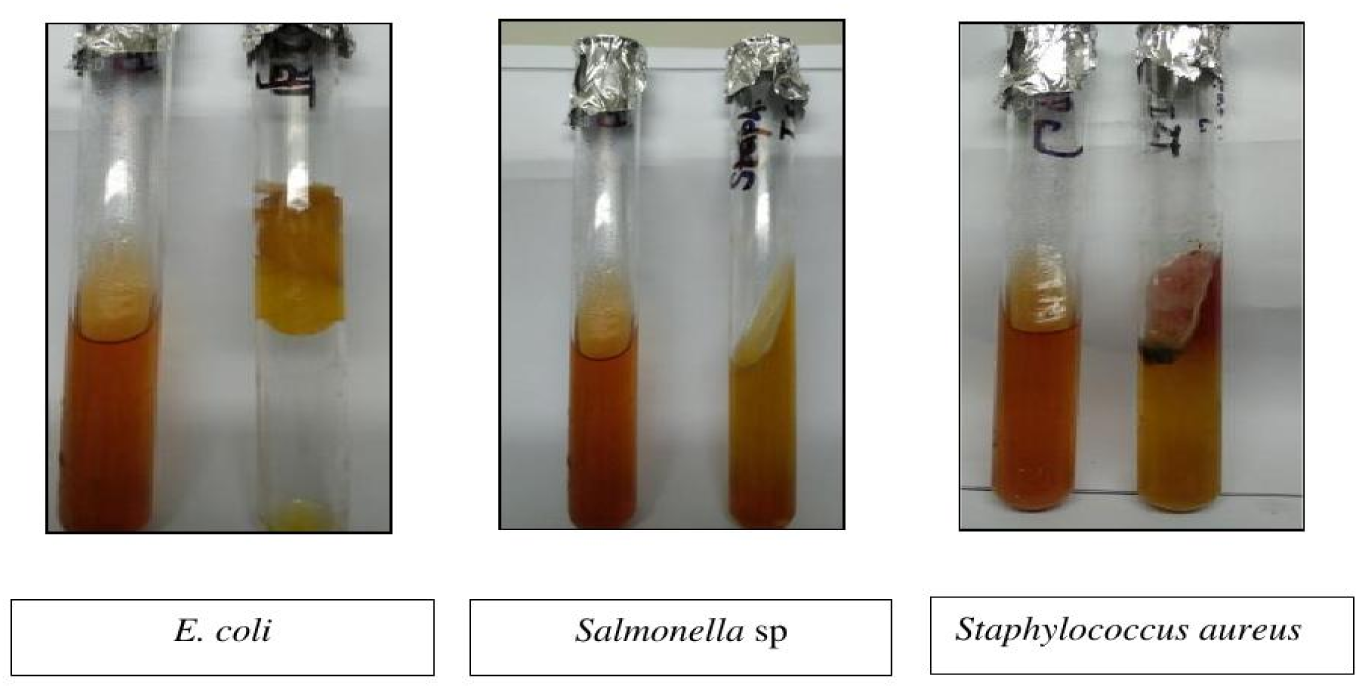
Isolated *E. coli*, *Salmonella* sp and *Staphylococcus aureus* were positive in TSI agar slant reaction. (left one is control)

### 3.3 Prevalence of bacteria in red meat

Of 60 samples examined, 6 (10.00%), 8 (13.33%) and 17 (28.33%) were positive overall for E. coli, Salmonella sp. and S. aureus, respectively. By district, the prevalence of E. coli, Salmonella sp. and S. aureus was 6.66%, 13.33% and 26.67% in Rajshahi, and 13.33%, 13.33% and 30.00% in Naogaon (Table 9).

**Table 9.** Overall prevalence of bacteria in red meat.

| Study area | Meat type | No. tested | E. coli positive (%) | Salmonella sp. positive (%) | S. aureus positive (%) |
| --- | --- | --- | --- | --- | --- |
| Rajshahi district | Cattle | 10 | 1 | 2 | 2 |
| Rajshahi district | Goat | 10 | 0 | 1 | 4 |
| Rajshahi district | Buffalo | 10 | 1 | 1 | 2 |
| Rajshahi total | — | 30 | 2 (6.66%) | 4 (13.33%) | 8 (26.67%) |
| Naogaon district | Cattle | 10 | 1 | 1 | 4 |
| Naogaon district | Goat | 10 | 1 | 1 | 3 |
| Naogaon district | Buffalo | 10 | 2 | 2 | 2 |
| Naogaon total | — | 30 | 4 (13.33%) | 4 (13.33%) | 9 (30.00%) |
| Grand total | — | 60 | 6 (10.00%) | 8 (13.33%) | 17 (28.33%) |

By meat type, the prevalence of E. coli, Salmonella sp. and S. aureus was 10.00%, 15.00% and 30.00% in cattle meat; 5.00%, 10.00% and 35.00% in goat meat; and 15.00%, 15.00% and 20.00% in buffalo meat, respectively (Table 10).

**Table 10.** Prevalence of bacteria in red meat according to meat type.

| Meat type | No. tested | E. coli (%) | Salmonella sp. (%) | S. aureus (%) |
| --- | --- | --- | --- | --- |
| Cattle meat | 20 | 2 (10.00%) | 3 (15.00%) | 6 (30.00%) |
| Goat meat | 20 | 1 (5.00%) | 2 (10.00%) | 7 (35.00%) |
| Buffalo meat | 20 | 3 (15.00%) | 3 (15.00%) | 4 (20.00%) |
| Total | 60 | 6 (10.00%) | 8 (13.33%) | 17 (28.33%) |

**Figure 18 & 19.** Motility test apparatus and antibiotic sensitivity pattern of isolated E. coli on Mueller-Hinton agar, showing sensitivity to gentamycin, ciprofloxacin and ceftriaxone, intermediate sensitivity to tetracycline, and resistance to penicillin, amoxicillin and ampicillin.

**Figure 18.**
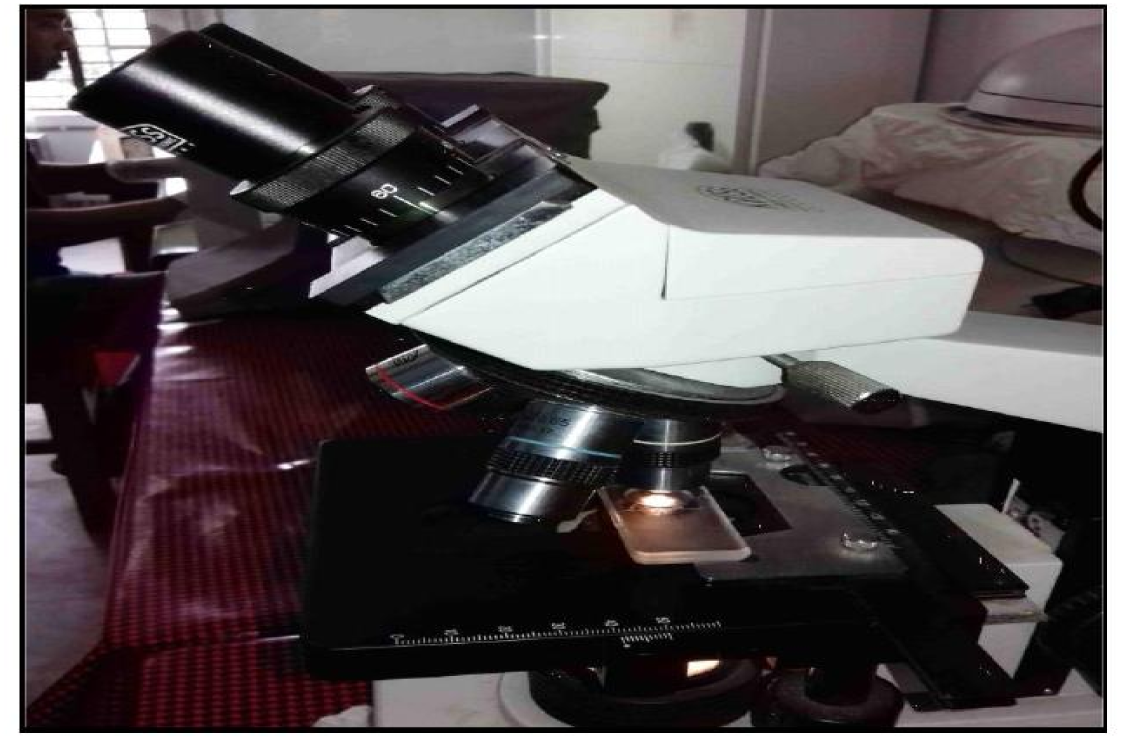
Isolated *E. coli* and *salmonella* sp showed motility in motility test.

**Figure 19.**
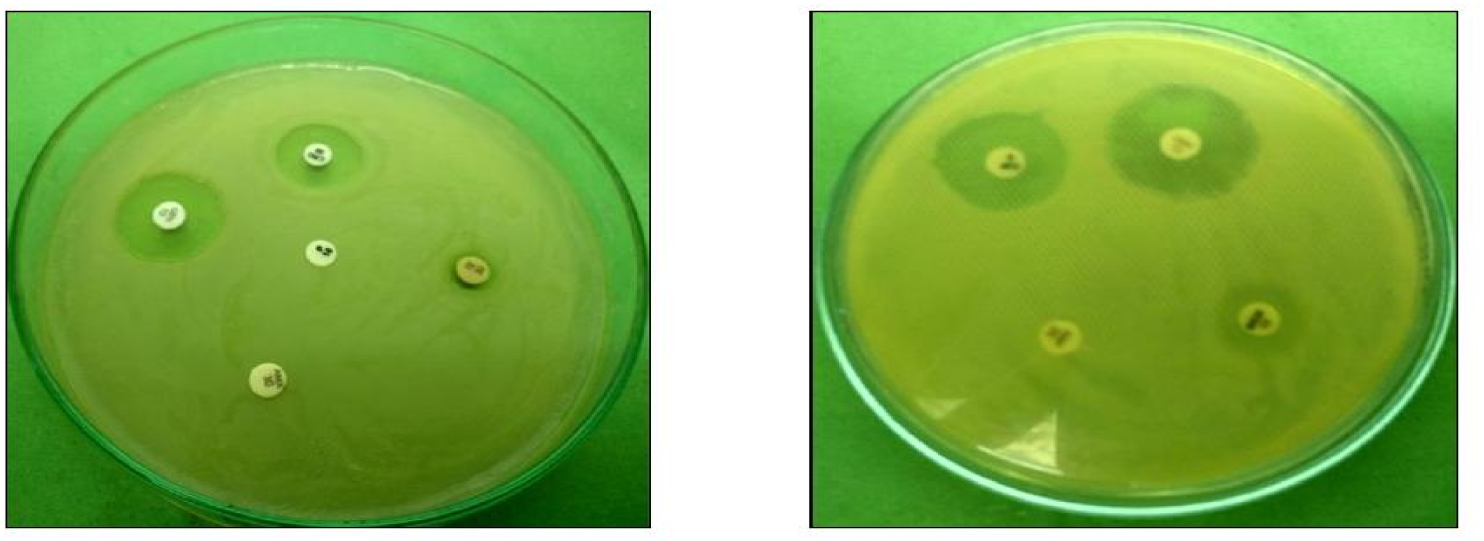
Antibiotic sensitivity pattern of isolated *E. coli* on Muller Hinton agar media showed sensitive to gentamycin and ciprofloxacin and ceftriaxone, intermediate sensitive to tetracycline but resistant to penicillin, amoxicillin and ampicillin.

### 3.4 Antibiotic sensitivity and resistance pattern of the isolates

E. coli isolates (n = 6) showed 100%, 83.33%, 50%, 33.33%, 16.67% and 16.67% resistance to penicillin, amoxicillin, ampicillin, tetracycline, ceftriaxone and gentamycin, respectively, with intermediate sensitivity of 66.67%, 33.33%, 16.67%, 16.67% and 16.67% to tetracycline, ceftriaxone, ampicillin, ciprofloxacin and gentamycin, respectively, and no intermediate sensitivity to penicillin or ampicillin was noted separately. Sensitivity was recorded as 83.33%, 66.67%, 50%, 16.67% and 16.67% to ciprofloxacin, gentamycin, ceftriaxone, amoxicillin and ampicillin, respectively, with no sensitivity to penicillin or tetracycline (Table 11).

**Table 11.** Antibiotic sensitivity and resistance pattern of E. coli isolated from red meat (n = 6)

| Antibiotic | Sensitive n (%) | Intermediate n (%) | Resistant n (%) |
| --- | --- | --- | --- |
| Penicillin | 0 (0%) | 0 (0%) | 6 (100%) |
| Amoxicillin | 1 (16.67%) | 0 (0%) | 5 (83.33%) |
| Ampicillin | 1 (16.67%) | 1 (16.67%) | 3 (50.0%) |
| Ceftriaxone | 3 (50.0%) | 2 (33.33%) | 1 (16.67%) |
| Ciprofloxacin | 5 (83.33%) | 1 (16.67%) | 1 (16.67%) |
| Gentamycin | 4 (66.67%) | 1 (16.67%) | 1 (16.67%) |
| Tetracycline | 0 (0%) | 4 (66.67%) | 2 (33.33%) |

Salmonella sp. isolates (n = 8) showed 87.5%, 75.0%, 25%, 25%, 12.5% and 12.5% resistance to penicillin, tetracycline, ampicillin, ceftriaxone, amoxicillin and ciprofloxacin, respectively, with no resistance to gentamycin. Sensitivity was recorded as 87.5%, 75%, 62.5%, 62.5%, 50% and 12.5% to gentamycin, ciprofloxacin, amoxicillin, ampicillin, ceftriaxone and tetracycline, respectively, with no sensitivity to penicillin (Table 12).

**Table 12.** Antibiotic sensitivity and resistance pattern of Salmonella sp. isolated from red meat (n = 8)

| Antibiotic | Sensitive n (%) | Intermediate n (%) | Resistant n (%) |
| --- | --- | --- | --- |
| Penicillin | 0 (0%) | 1 (12.5%) | 7 (87.5%) |
| Amoxicillin | 5 (62.5%) | 2 (25.0%) | 1 (12.5%) |
| Ampicillin | 5 (62.5%) | 1 (12.5%) | 2 (25.0%) |
| Ceftriaxone | 4 (50.0%) | 2 (25.0%) | 2 (25.0%) |
| Ciprofloxacin | 6 (75.0%) | 1 (12.5%) | 1 (12.5%) |
| Gentamycin | 7 (87.5%) | 1 (12.5%) | 0 (0%) |
| Tetracycline | 1 (12.5%) | 1 (12.5%) | 6 (75.0%) |

S. aureus isolates (n = 17) showed 94.11%, 58.82%, 47.05%, 47.05%, 17.65%, 11.76% and 11.76% resistance to penicillin, tetracycline, ampicillin, amoxicillin, ciprofloxacin, ceftriaxone and gentamycin, respectively, with intermediate sensitivity of 35.29%, 23.52%, 17.64%, 11.76%, 5.89% and 5.89% to tetracycline, ciprofloxacin, ceftriaxone, amoxicillin, ampicillin and penicillin, respectively, and no intermediate sensitivity to gentamycin. Sensitivity was recorded as 88.24%, 75%, 70.59%, 41.18%, 35.29% and 17.65% to gentamycin, ceftriaxone, ciprofloxacin, amoxicillin, ampicillin and tetracycline, respectively, with no sensitivity to penicillin (Table 13).

**Table 13.** Antibiotic sensitivity and resistance pattern of Staphylococcus aureus isolated from red meat (n = 17)

| Antibiotic | Sensitive n (%) | Intermediate n (%) | Resistant n (%) |
| --- | --- | --- | --- |
| Penicillin | 0 (0%) | 1 (5.89%) | 16 (94.11%) |
| Amoxicillin | 7 (41.18%) | 2 (11.76%) | 8 (47.05%) |
| Ampicillin | 6 (35.29%) | 1 (5.89%) | 10 (58.82%) |
| Ceftriaxone | 12 (70.59%) | 3 (17.64%) | 2 (11.76%) |
| Ciprofloxacin | 11 (75.0%) | 4 (23.52%) | 3 (17.65%) |
| Gentamycin | 15 (88.24%) | 0 (0%) | 2 (11.76%) |
| Tetracycline | 3 (17.65%) | 6 (35.29%) | 8 (47.05%) |

**Figure 20 & 21.** Antibiotic sensitivity pattern of isolated Salmonella sp. (sensitive to ciprofloxacin and gentamycin; intermediate to tetracycline; resistant to ampicillin, amoxicillin and penicillin) and Staphylococcus aureus (sensitive to gentamycin and ceftriaxone; intermediate to tetracycline; resistant to penicillin, ampicillin and amoxicillin) on Mueller-Hinton agar.

**Figure 20.**
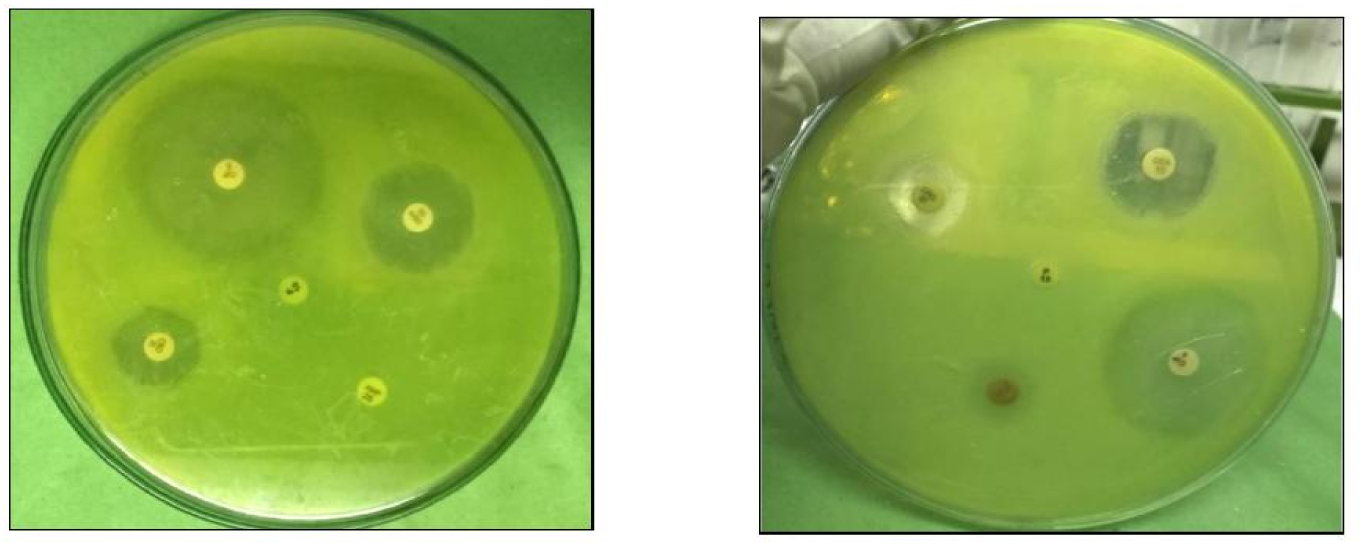
Antibiotic sensitivity pattern of isolated *Salmonella* sp on Muller Hinton agar media showed sensitive to ciprofloxacin, gentamycin but intermediate sensitive to Tetracycline but resistant to ampicillin, amoxacilin and penicillin.

**Figure 21.**
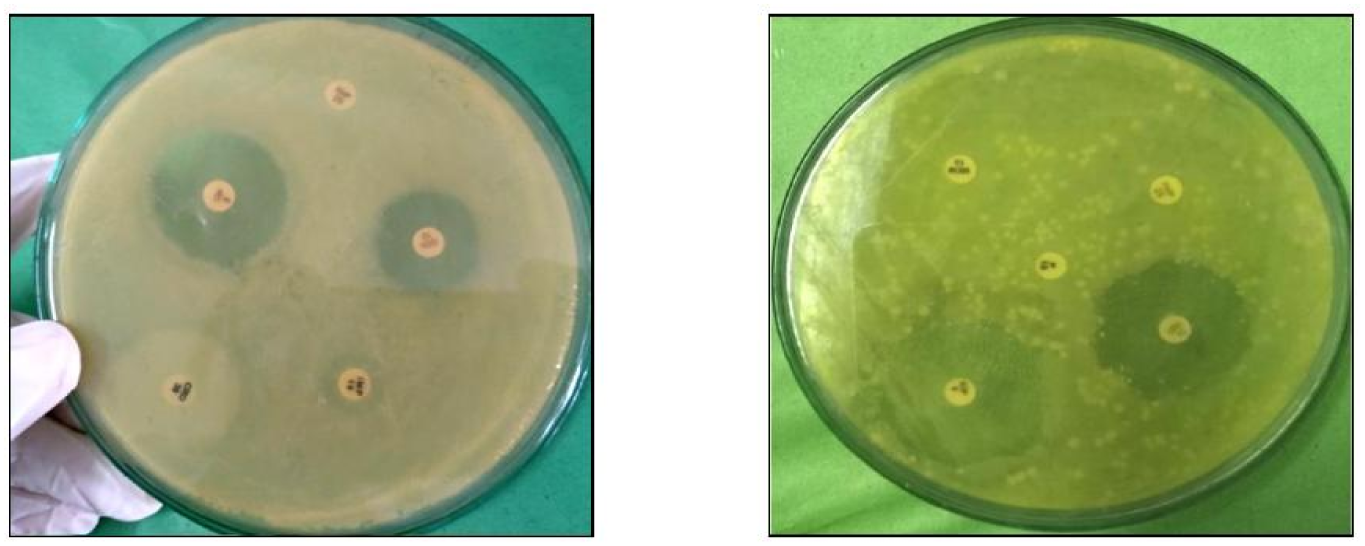
Antibiotic sensitivity pattern of isolated *Staphylococcus aureus* on Muller Hinton agar media showed sensitive to gentamycin, ceftriaxone but intermediate sensitive to Tetracycline but resistant to penicillin, ampicillin and amoxicillin.

### 3.5 Detection of antibiotic residues

Fifteen samples (five cattle, five goat and five buffalo) were screened for antibiotic residues by TLC. Of these, two cattle-meat samples were positive: one (6.67%) for ciprofloxacin and one (6.67%) for oxytetracycline. No residues were detected for penicillin, ampicillin, amoxicillin, ceftriaxone, gentamycin, streptomycin, cloxacillin or sulphonamides, and no residues were detected in goat or buffalo meat (Table 14).

**Table 14.** Detection of antibiotic residues in red meat samples (n = 15)

| Antibiotic | Residue-positive samples, n (%) |
| --- | --- |
| Penicillin | 0 (0%) |
| Ciprofloxacin | 1 (6.67%) |
| Ampicillin | 0 (0%) |
| Oxytetracycline | 1 (6.67%) |
| Amoxicillin | 0 (0%) |
| Ceftriaxone | 0 (0%) |
| Gentamycin | 0 (0%) |
| Streptomycin | 0 (0%) |
| Cloxacillin | 0 (0%) |
| Sulphonamides | 0 (0%) |

## 4. Discussion

Meat is an important source of protein and a valuable resource in poor communities; however, as a rich nutrient source it can also be a vehicle for human food-borne illness, and inappropriate slaughtering and retail practices can compromise food safety, particularly in densely populated settlements (Datt et al., 2003). In the current study, the cultural, staining and biochemical characteristics of the isolated E. coli, Salmonella sp. and S. aureus were consistent with the findings of Freeman (1985), Buxton and Fraser (1977) and Merchant and Packer (1967).

The prevalence of E. coli in the present study (10.00%) is in agreement with the findings of Shekh et al. (2013), who isolated 8.80% E. coli from buffalo meat sold in Parbhani city, Maharashtra, India; Iroha et al. (2011), who reported 8.00% E. coli in raw meat sold in retail shops in Abakaliki, Ebonyi State, Nigeria; and Jahan et al. (2015), who isolated 10.00% E. coli from fresh raw beef sold in markets of Sylhet Sadar, Bangladesh. The present result is markedly lower than that of Gayathri and Anu (2015), who isolated 70% E. coli from raw meat sold in different markets around Chennai, India, but somewhat higher than the 37.50% reported by Seran et al. (2012) in Turkey. Such variation in prevalence may reflect differences in hygienic management of the slaughtering area, ambient temperature and other sources of environmental contamination.

In this study, 13.33% of the examined samples were contaminated with Salmonella sp., a finding supported by Jahan et al. (2015), who reported 13.33% Salmonella sp. from fresh raw beef in Sylhet Sadar, Bangladesh, and broadly consistent with the 15% reported by Gayathri and Anu (2015) in Chennai, India. However, Hasan et al. (2018) reported a considerably higher prevalence of 46.67% Salmonella sp. in meat from different districts of Bangladesh, and Victoria and Tajudeen (2011) isolated Salmonella sp. from 50% of retail meat samples in Ibadan municipal abattoir, Nigeria. S. aureus was recovered from 28.33% of the examined samples, closely comparable with the findings of Jahan et al. (2015) (26.67%) and Gayathri and Anu (2015) (25%). By contrast, Hanston et al. (2011) reported a lower prevalence of 17.8% S. aureus on retail meat in Iowa, and Iroha et al. (2011) reported only 1.3% contamination in Abakaliki, Nigeria. Variation in reported prevalence of S. aureus may reflect differences in sample origin, sample-collection and transportation technique, and prevailing environmental conditions.

With respect to antimicrobial susceptibility, the resistance profile of E. coli isolates recorded in the present study is broadly consistent with the findings of Rahman et al. (2017), who reported that 85.71% and 71.43% of E. coli isolated from beef were resistant to erythromycin and oxytetracycline, respectively, and 100% sensitive to ciprofloxacin, gentamicin and neomycin. Kamana et al. (2019) similarly reported that E. coli exhibited the highest resistance to amoxicillin (100%), followed by tetracycline (93%), nalidixic acid (25%) and cefotaxime (19%) in meat samples from eastern Nepal, whereas Tricia et al. (2006) reported 43% resistance to ampicillin among E. coli isolates but no resistance to gentamicin.

The resistance pattern of Salmonella sp. observed here is consistent with the findings of Hasan et al. (2018), who reported that all Salmonella isolates were resistant to amoxicillin, and with those of Kamana et al. (2019), who reported the highest resistance to amoxicillin (100%), followed by tetracycline (24%), chloramphenicol (11%) and nalidixic acid (11%) in isolates from chicken, pork, goat and buffalo meat in eastern Nepal. Victoria and Tajudeen (2011) also reported that Salmonella sp. isolated from retail meat was highly sensitive to gentamicin and amoxicillin but resistant to tetracycline, ciprofloxacin, ampicillin and norfloxacin.

The resistance profile of S. aureus recorded in this study is broadly similar to that reported by Kamana et al. (2019), who found S. aureus resistant to amoxicillin (100%), tetracycline (63%) and cefotaxime (13%) in meat from eastern Nepal, and by Feben et al. (2018), who reported that 49.5%, 45.5%, 45% and 13% of S. aureus strains isolated from beef in Ethiopia were resistant to penicillin G, vancomycin, cloxacillin and norfloxacin, respectively, while 86.5%, 73%, 72%, 54% and 50% were susceptible to amoxicillin, norfloxacin, erythromycin, cloxacillin and penicillin G, respectively. Variation in sensitivity patterns across studies may reflect differing degrees of prior antibiotic exposure and indiscriminate use of antimicrobials as feed additives or as preventive or curative agents.

The detection of antibiotic residues by TLC (6.67% positive for ciprofloxacin and 6.67% for oxytetracycline) is lower than several previously reported values. Tsepo et al. (2017) reported that sulphanilamide had the highest positivity (92.5%), followed by streptomycin (29.4%), ciprofloxacin (21.4%) and tetracycline (14.6%), while Bilatu (2012) reported 28% tetracycline, 23% sulfonamide and 20% penicillin residues in fresh and ready-to-eat beef in Thailand, and Shaltout et al. (2019) reported considerably higher residue rates of ciprofloxacin (20.74%) and oxytetracycline (87.97%) in fresh beef marketed at Giza governorate, Egypt. Variation in reported residue levels across studies may reflect differences in the extent of antibiotic use and adherence to the recommended withdrawal period following treatment of food animals.

## 5. Conclusion

This study demonstrates that retail red meat marketed in Rajshahi and Naogaon districts is contaminated with multidrug-resistant Escherichia coli, Salmonella spp., and Staphylococcus aureus, with detectable residues of ciprofloxacin and oxytetracycline in a proportion of samples. These findings highlight potential risks to food safety and public health through the dissemination of antimicrobial-resistant bacteria and antibiotic residues via the food chain. Strengthening hygienic slaughtering and meat-handling practices, promoting prudent antimicrobial use in food-producing animals, ensuring compliance with drug withdrawal periods, and implementing routine surveillance programs are essential to reduce contamination and mitigate the spread of antimicrobial resistance.

## Acknowledgements

The authors gratefully acknowledge the Department of Veterinary & Animal Sciences, University of Rajshahi, Bangladesh, for providing laboratory facilities and technical support throughout this study. The authors also sincerely thank the retail meat vendors and local authorities of Rajshahi and Naogaon districts for their cooperation during sample collection.

## Ethical Approval

Ethical approval for this study was obtained from the Institutional Animal, Medical Ethics, Biosafety and Biosecurity Committee (IAMEBBC), University of Rajshahi, Bangladesh, before the commencement of the study. Meat samples were collected from retail meat shops with the permission of the respective vendors. No live animals or human participants were directly involved in this study.

## Informed Consent

Permission was obtained from the respective retail meat vendors prior to sample collection. All authors have read and approved the final version of the manuscript and agree to its submission for publication.

## Data Availability

The data supporting the findings of this study are available from the corresponding author upon reasonable request. All data generated or analyzed during this study are included in this article or are available from the corresponding author upon reasonable request.

## Author Contributions

Nise Saha: Conceptualization, Literature review, Sample collection, Data curation, Laboratory experiments, Data analysis, Methodology, Statistical analysis, Writing-original draft.

Sharmin afroz: Sample collection, Data curation, Literature review, Laboratory experiments, Investigation, Data analysis, Writing-original draft

Md. Rimon Bhuiyan: Investigation, Data curation, Data analysis, Laboratory experiments, Visualization, Writing-original draft, Writing-review and editing.

Anna Purnna Ray: Data curation, Validation, Investigation, Data analysis, Writing-original draft.

Kallyanmoy Das, Md. Ahsan Hasan Jony, Rubina Khatun: Data curation, Validation, Investigation.

K.M. Mozaffor Hossain: Conceptualization, Methodology, Supervision, Project administration, Writing-review and editing, Final approval of the manuscript.

## Funding

This research received no specific grant from any funding agency in the public, commercial, or not-for-profit sectors.

## Conflict of Interest

The authors declare that they have no competing interests and no conflict of interest regarding the publication of this manuscript.

## Notes

### Competing Interest Statement

The authors have declared no competing interest.

